# The single item burnout measure is a reliable and valid tool to measure occupational burnout

**DOI:** 10.1101/2023.03.06.23286842

**Authors:** P. Galanis, A. Katsiroumpa, I. Vraka, O. Siskou, O. Konstantakopoulou, T. Katsoulas, P. Gallos, Daphne Kaitelidou

## Abstract

**OBJECTIVE:** To estimate the reliability and the validity of the single item burnout measure in a sample of nurses in Greece.

**METHOD:** We conducted an online cross-sectional study in Greece with 963 nurses. Data were collected during October 2022. We measured demographic and work-related variables of nurses, i.e. gender, age, chronic disease, self-rated health status, years of experience, and working in COVID-19 ward/intensive care unit. We used the single item burnout (SIB) and the Copenhagen Burnout Inventory (CBI) to measure occupational burnout. Moreover, we used the COVID-19 burnout scale (COVID-19-BS) to measure nurses’ burnout during the pandemic, and the Patient Health Questionnaire-4 (PHQ-4) to measure anxiety and depression among nurses.

**RESULTS:** Intraclass correlation coefficient between the two measurements of the SIB during the test-retest study was 0.986 indicating excellent reliability of the SIB. We found a high correlation between CBI factors and SIB (p<0.001), a moderate correlation between PHQ-4 and SIB (p<0.001), and a low to moderate correlation between COVID-19-BS and SIB (p<0.001). Therefore, concurrent validity of SIB was excellent. Moreover, SIB had high discriminant validity. In particular, nurses with a chronic disease, those with a very poor/poor/moderate health status, and those working in COVID-19 ward/intensive care unit had higher levels of burnout according to the SIB (p<0.001 in all cases). Moreover, we found a positive relationship between years of experience and SIB score (r=0.13, p<0.001).

**CONCLUSIONS:** The single item burnout measure is a brief, reliable, and valid tool that we can use as a screening measure to identify individuals at high risk of burnout.

## Introduction

The World Health Organization defines occupational burnout an occupational phenomenon caused by chronic stress due to work or the workplace.^1^ Occupational burnout is not considered as a medical condition and occurs when individuals cannot manage effectively their chronic stress. Burnout is mainly characterized by exhaustion, cynicism, and inefficacy.^2^ In particular, people that suffer from burnout are also emotionally exhausted, show increased levels of depersonalisation and cynicism, and feel a reduced personal accomplishment.^3^

Burnout is prevalent in a variety of jobs. In particular, the pooled prevalence of burnout in 43.2% in physicians,^4^ in nurses is 11.2%,^5^ in pharmacists is 51%,^6^ in general practitioners is 37%,^7^ in dentists is 13%,^8^ in psychiatrists is 25.9%,^9^ and in primary healthcare professionals is 28.1%^10^. We should notice that there are significant differences in prevalence of burnout between geographical regions, clinical settings and specialties. COVID-19 pandemic has had a tremendous impact on physical and mental health of healthcare workers increasing their burnout.^11–13^ High levels of burnout among healthcare workers is an occupational hazard since burnout is related with decreased healthcare workers productivity and patients satisfaction, and worsening quality of care and safety.^14,15^

Several questionnaires, tools, and scales are available for measuring burnout, such as the Maslach Burnout Inventory, the Oldenburg Burnout Inventory, the Copenhagen Burnout Inventory, the Shirom-Melamed Burnout Measure, and the Burnout Clinical Subtype Questionnaire.^16^ Among these tools, the Maslach Burnout Inventory and the Copenhagen Burnout Inventory are the most widely used tools in healthcare research to measure burnout. The main disadvantage of these tools is that they consist of many items causing participants’ tiredness and low response rates. Thus, a single item burnout measure is created in order to measure occupational burnout quickly and valid.^17^ The aim of our study was to estimate the reliability and the validity of the single item burnout measure in a sample of nurses in Greece.

## Methods

### Study design

We conducted an online cross-sectional study in Greece with 963 nurses. Data were collected during October 2022. We created an online version of the study questionnaire and we disseminated it through social media. Thus, a convenience sample with unknown response rate was obtained. We applied the following inclusion criteria: (a) adults participants, (b) working as nurses, (c) and participants who understand the Greek language. Prior to the final study, we conducted a pilot study with 50 nurses in order to perform the test-retest method. In that case, nurses completed two times the questionnaire with an interval of one week. Moreover, we performed cognitive interviews with ten nurses in order to assess the face validity of the questionnaire. Face validity was excellent since all nurses understand and complete the study questionnaire.

We collected our data in an anonymous and voluntary basis. Moreover, we informed participants about the aim and the design of the study and they gave their informed consent. In addition, our study protocol was approved by the Ethics Committee of Faculty of Nursing, National and Kapodistrian University of Athens (reference number; 417, 7 September 2022). Also, we followed the guidelines of the Declaration of Helsinki in order to conduct our study.

### Measurements

We measured demographic and work-related variables of nurses, i.e. gender (females or males), age (continuous variable), chronic disease (no or yes), self-rated health status (scale from 1 [very poor] to 5 [very good]), years of experience (continuous variable), and working in COVID-19 ward/intensive care unit (no or yes).

We used the single item burnout (SIB) to measure occupational burnout.^17^ In that case, we asked nurses to rate their current level of burnout. In particular, the question was the following: “In a scale from 0 (not at all) to 10 (totally), how tired do you feel?”.

Moreover, we used the Copenhagen Burnout Inventory (CBI) to measure occupational burnout.^18^ The CBI consists of 19 items creating three factors: personal burnout, work-related burnout, and client-related burnout. Score on the three factors ranges from zero (not at all burnout) to 100 (extreme burnout). We used the Greek version of the CBI which is proven to be reliable and valid.^19^

Also, we used the COVID-19 burnout scale (COVID-19-BS) to measure nurses’ burnout since we performed our study three years after the onset of the COVID-19 pandemic and COVID-19 burnout among nurses was possible.^20^ The COVID-19-BS includes 13 items creating three factors: emotional exhaustion, physical exhaustion, and exhaustion due to measures against the COVID-19. Score on the three factors ranges from one (not at all burnout) to five (extreme burnout). We used the reliable and valid Greek version of the COVID-19-BS.^20,21^

We used the Patient Health Questionnaire-4 (PHQ-4) to measure anxiety and depression among nurses.^22^ Two items measure the anxiety and two items measure the depression creating a score from 0 (normal levels) to 6 (severe symptomatology). Greek version of the PHQ-4 seems to be reliable and valid.^23^

### Statistical analysis

We use numbers and percentages to present categorical variables, and means and standard deviations to present continuous variables. We calculated intraclass correlation coefficient between the two measurements of the SIB during the test-retest study. We calculated Pearson’s correlation coefficient between the SIB and CBI, COVID-19-BS, and PHQ-4 in order to estimate the concurrent validity of the SIB. Also, we conducted known-group analysis by performing the following: (a) independent samples t-test for gender, chronic disease, health status, and working in COVID-19 ward/intensive care unit, (b) Pearson’s correlation coefficient for age, and (c) Spearman’s correlation coefficient for years of experience. P-values less than 0.05 were considered as statistically significant. We used the IBM SPSS 21.0 (IBM Corp. Released 2012. IBM SPSS Statistics for Windows, Version 21.0. Armonk, NY: IBM Corp.) for the analysis.

## Results

Study population included 963 nurses. Most of nurses were females (88.4%) in a good/very good health (88.4%). Mean age was 37.9 years, while mean years of experience was 12. One out of four nurses (25%) reported a chronic disease, while 64.1% working in COVID-19 ward/intensive care unit. Detailed demographic and work-related data of nurses are presented in Table 1.

**Table 1.** Demographic and work-related data of nurses.

| Variables | N | % |
| --- | --- | --- |
| Gender |  |  |
| Males | 112 | 11.6 |
| Females | 851 | 88.4 |
| Age (years)<sup>a</sup> | 37.9 | 9.6 |
| Chronic disease |  |  |
| No | 722 | 75.0 |
| Yes | 241 | 25.0 |
| Self-perceived health status |  |  |
| Very poor/poor/moderate | 112 | 11.6 |
| Good/very good | 851 | 88.4 |
| Working in COVID-19 ward/intensive care unit |  |  |
| No | 346 | 35.9 |
| Yes | 617 | 64.1 |
| Years of experience<sup>a</sup> | 12.0 | 9.2 |

<sup>a</sup> mean, standard deviation

Intraclass correlation coefficient between the two measurements of the SIB during the test-retest study was 0.986 (95% confidence interval = 0.976 to 0.992, p<0.001) indicating excellent reliability of the SIB.

Correlations between SIB and the other scales are presented in Table 2. All correlations were statistically significant (p<0.001 in all cases) and therefore concurrent validity of the SIB was excellent. In particular, we found a high correlation between Copenhagen Burnout Inventory factors and the SIB, a moderate correlation between the Patient Health Questionnaire-4 and the SIB, and a low to moderate correlation between the COVID-19 burnout scale and the SIB.

**Table 2.** Correlations between the single item burnout measure and the Copenhagen Burnout Inventory, the COVID-19 burnout scale, and the Patient Health Questionnaire-4

| Scale | Single item burnout |  |
| --- | --- | --- |
|  | Correlation coefficient | P-value |
| Copenhagen Burnout Inventory |  |  |
| Personal burnout | 0.82 | <0.001 |
| Work-related burnout | 0.72 | <0.001 |
| Client-related burnout | 0.79 | <0.001 |
| COVID-19 burnout scale |  |  |
| Emotional exhaustion | 0.45 | <0.001 |
| Physical exhaustion | 0.53 | <0.001 |
| Exhaustion due to measures against the COVID-19 | 0.21 | <0.001 |
| Patient Health Questionnaire-4 |  |  |
| Anxiety | 0.42 | <0.001 |
| Depression | 0.46 | <0.001 |

Results from known-groups analysis are presented in Table 3. SIB had high discriminant validity. In particular, nurses with a chronic disease, those with a very poor/poor/moderate health status, and those working in COVID-19 ward/intensive care unit had higher levels of burnout according to the SIB (p<0.001 in all cases). Moreover, we found a positive relationship between years of experience and SIB score (r=0.13, p<0.001).

**Table 3.** Known-groups analysis between the single item burnout measure and demographic and work-related data of nurses.

| Variables | Single item burnout measure |  | P-value |
| --- | --- | --- | --- |
|  | Mean | Standard deviation |  |
| Gender |  |  | 0.48<sup>a</sup> |
| Males | 6.32 | 2.57 |  |
| Females | 6.51 | 2.58 |  |
| Age (years) |  | 0.05<sup>b</sup> | 0.14<sup>b</sup> |
| Chronic disease |  |  | <0.001<sup>a</sup> |
| No | 6.24 | 2.62 |  |
| Yes | 7.20 | 2.31 |  |
| Self-perceived health status |  |  | <0.001<sup>a</sup> |
| Very poor/poor/moderate | 6.76 | 2.44 |  |
| Good/very good | 5.79 | 2.78 |  |
| Working in COVID-19 ward/intensive care unit |  |  | <0.001<sup>a</sup> |
| No | 6.02 | 2.55 |  |
| Yes | 6.75 | 2.56 |  |
| Years of experience |  | 0.13<sup>c</sup> | <0.001<sup>c</sup> |

<sup>a</sup> independent samples t-test
<sup>b</sup> Pearson's correlation coefficient
<sup>c</sup> Spearman's correlation coefficient

## Discussion

We conducted a cross-sectional study to assess the psychometric properties of the single item burnout measure. We found that the single item burnout measure is a reliable and valid tool that we can use to measure occupational burnout easy and fast.

A brief and sensitive tool as SIB is imperative to identify workers burnout since this occupational phenomenon is related with physical and mental health, and turnover intention. Our results support the hypothesis that SIB can fulfill this gap due its reliability, validity, ease of administration, and brevity. First, we found that the SIB had excellent reliability in our pilot study performing the test-retest method. Moreover, concurrent validity and known-groups analysis confirmed the high level of validity of SIB. We used three other scales (i.e., Copenhagen Burnout Inventory, Patient Health Questionnaire-4, COVID-19 burnout scale) to measure concurrent validity of the SIB and six demographic and work-related data of nurses to measure discriminant validity.

Our finding are confirmed by several other studies that estimate the psychometric properties of the SIB.^24–26^ These studies used the Maslach Burnout Inventory as a gold standard to compare the SIB, while the study populations included general practitioners and primary care staff. Scholars found that SIB is a sensitive and specific tool to identify workers at high or low levels of burnout with a high degree of accuracy.

Literature supports our results from the known-groups analysis. In particular, a recent systematic review confirms that healthcare professionals working with COVID-19 patients are more likely to experience burnout, stress, and depression.^27^ Moreover, we found that nurses with a very poor/poor/moderate health status and those with a chronic disease experienced higher level of burnout. This finding is suggested from previous research where healthcare workers who suffered from several diseases such as depression, anxiety, stress.^28–30^

Our study had several limitations. First, we assessed reliability and validity of the SIB using different methods and tools. However, other analyses such as sensitivity and specificity analysis could be performed in order to get more valid results. Also, other tools such as Maslach Burnout Inventory could be used as gold standard in order to compare the SIB with them. Moreover, we used a big sample of nurses but further studies with different professionals (e.g. physicians, workers in primary care services, dentists, etc.) should be conducted in order to expand our results. Additionally, we performed known-groups analysis using six demographic and work-related data of nurses. Future research could use more demographic and work-related variables in order to investigate the discriminant validity of the SIB.

In conclusion, the single item burnout measure is a reliable and valid tool that we can use to measure occupational burnout. Since burnout among healthcare workers is highly prevalent tools like the SIB could be used as sensitive and brief screening measures to identify individuals at high risk of burnout.

## Data Availability

All data produced in the present study are available upon reasonable request to the authors

## References

1. World Health Organization. Burn-out an “occupational phenomenon”: International Classification of Diseases [Internet]. 2019. Available from: https://www.who.int/news/item/28-05-2019-burn-out-an-occupational-phenomenon-international-classification-of-diseases

2. Maslach C, Schaufeli WB, Leiter MP. Job burnout. Annu Rev Psychol. 2001;52:397–422.

3. Bianchi R, Schonfeld IS, Laurent E. Is it time to consider the “burnout syndrome” a distinct illness? Frontiers in Public Health. 2015 Jun 8;3:158.

4. Hiver C, Villa A, Bellagamba G, Lehucher-Michel MP. Burnout prevalence among European physicians: a systematic review and meta-analysis. Int Arch Occup Environ Health. 2022 Jan;95(1):259–73.

5. Woo T, Ho R, Tang A, Tam W. Global prevalence of burnout symptoms among nurses: A systematic review and meta-analysis. J Psychiatr Res. 2020 Apr;123:9–20.

6. Dee J, Dhuhaibawi N, Hayden JC. A systematic review and pooled prevalence of burnout in pharmacists. Int J Clin Pharm. 2022 Nov 29;1–10.

7. Shen X, Xu H, Feng J, Ye J, Lu Z, Gan Y. The global prevalence of burnout among general practitioners: a systematic review and meta-analysis. Fam Pract. 2022 Sep 24;39(5):943–50.

8. Moro J da S, Soares JP, Massignan C, Oliveira LB, Ribeiro DM, Cardoso M, et al. Burnout syndrome among dentists: A systematic review and meta-analysis. J Evid Based Dent Pract. 2022 Sep;22(3):101724.

9. Bykov KV, Zrazhevskaya IA, Topka EO, Peshkin VN, Dobrovolsky AP, Isaev RN, et al. Prevalence of burnout among psychiatrists: A systematic review and meta-analysis. J Affect Disord. 2022 Jul 1;308:47–64.

10. Wright T, Mughal F, Babatunde OO, Dikomitis L, Mallen CD, Helliwell T. Burnout among primary health-care professionals in low- and middle-income countries: systematic review and meta-analysis. Bull World Health Organ. 2022 Jun 1;100(6):385–401A.

11. Galanis P, Vraka I, Fragkou D, Bilali A, Kaitelidou D. Nurses’ burnout and associated risk factors during the COVID-19 pandemic: A systematic review and meta-analysis. J Adv Nurs. 2021 Aug;77(8):3286–302.

12. Ghahramani S, Lankarani KB, Yousefi M, Heydari K, Shahabi S, Azmand S. A Systematic Review and Meta-Analysis of Burnout Among Healthcare Workers During COVID-19. Front Psychiatry. 2021;12:758849.

13. Macaron MM, Segun-Omosehin OA, Matar RH, Beran A, Nakanishi H, Than CA, et al. A systematic review and meta analysis on burnout in physicians during the COVID-19 pandemic: A hidden healthcare crisis. Front Psychiatry. 2022;13:1071397.

14. Jun J, Ojemeni MM, Kalamani R, Tong J, Crecelius ML. Relationship between nurse burnout, patient and organizational outcomes: Systematic review. Int J Nurs Stud. 2021 Jul;119:103933.

15. Mossburg SE, Dennison Himmelfarb C. The Association Between Professional Burnout and Engagement With Patient Safety Culture and Outcomes: A Systematic Review. J Patient Saf. 2021 Dec 1;17(8):e1307–19.

16. Alahmari MA, Al Moaleem MM, Hamdi BA, Hamzi MA, Aljadaani AT, Khormi FA, et al. Prevalence of Burnout in Healthcare Specialties: A Systematic Review Using Copenhagen and Maslach Burnout Inventories. Med Sci Monit [Internet]. 2022 Dec 7 [cited 2023 Mar 5];28. Available from: https://www.medscimonit.com/abstract/index/idArt/938798

17. Hansen V, Pit S. The Single Item Burnout Measure is a Psychometrically Sound Screening Tool for Occupational Burnout. Health Scope [Internet]. 2016 Jan 3 [cited 2023 Mar 5];5(2). Available from: https://brieflands.com/articles/healthscope-20154.html

18. Kristensen TS, Borritz M, Villadsen E, Christensen KB. The Copenhagen Burnout Inventory: A new tool for the assessment of burnout. Work & Stress. 2005 Jul;19(3):192–207.

19. Papaefstathiou E, Tsounis A, Malliarou M, Sarafis P. Translation and validation of the Copenhagen Burnout Inventory amongst Greek doctors. Health Psych Res [Internet]. 2019 Sep 18 [cited 2023 Mar 5];7(1). Available from: https://healthpsychologyresearch.openmedicalpublishing.org/article/22444

20. Galanis P, Katsiroumpa A, Sourtzi P, Siskou O, Konstantakopoulou O, Kaitelidou D. The COVID-19 burnout scale: Development and initial validation [Internet]. 2022 [cited 2022 Nov 19]. Available from: http://medrxiv.org/lookup/doi/10.1101/2022.10.20.22281317

21. Galanis P, Katsiroumpa A, Sourtzi P, Siskou O, Konstantakopoulou O, Katsoulas T, et al. COVID-19-Related Burnout and Intention of Fully Vaccinated Individuals to Get a Booster Dose: The Mediating Role of Resilience. Vaccines. 2022 Dec 27;11(1):62.

22. Kroenke K, Spitzer RL, Williams JBW, Lowe B. An Ultra-Brief Screening Scale for Anxiety and Depression: The PHQ-4. Psychosomatics. 2009 Nov 1;50(6):613–21.

23. Karekla M, Pilipenko N, Feldman J. Patient Health Questionnaire: Greek language validation and subscale factor structure. Comprehensive Psychiatry. 2012 Nov;53(8):1217–26.

24. Hansen V, Girgis A. Can a single question effectively screen for burnout in Australian cancer care workers? BMC Health Serv Res. 2010 Dec;10(1):341.

25. Rohland BM, Kruse GR, Rohrer JE. Validation of a single-item measure of burnout against the Maslach Burnout Inventory among physicians. Stress and Health. 2004 Apr;20(2):75–9.

26. Dolan ED, Mohr D, Lempa M, Joos S, Fihn SD, Nelson KM, et al. Using a Single Item to Measure Burnout in Primary Care Staff: A Psychometric Evaluation. J GEN INTERN MED. 2015 May;30(5):582–7.

27. Ulfa M, Azuma M, Steiner A. Burnout status of healthcare workers in the world during the peak period of the COVID-19 pandemic. Front Psychol. 2022;13:952783.

28. Chen C, Meier ST. Burnout and depression in nurses: A systematic review and meta-analysis. Int J Nurs Stud. 2021 Dec;124:104099.

29. Kratzke IM, Woods LC, Adapa K, Kapadia MR, Mazur L. The Sociotechnical Factors Associated With Burnout in Residents in Surgical Specialties: A Qualitative Systematic Review. J Surg Educ. 2022;79(3):614–23.

30. Meredith LS, Bouskill K, Chang J, Larkin J, Motala A, Hempel S. Predictors of burnout among US healthcare providers: a systematic review. BMJ Open. 2022 Aug 25;12(8):e054243.

